# SEPRES: Sepsis prediction via the clinical data integration system in the ICU

**DOI:** 10.1101/2022.07.06.22277188

**Authors:** Qiyu Chen, Ranran Li, ChihChe Lin, Chiming Lai, Yaling Huang, Wenlian Lu, Lei Li

## Abstract

**Background:** The lack of information interoperability between different devices and systems in the ICU hinders further utilization of data, especially for early warning of specific diseases in the ICU.

**Objectives:** We aimed to establish a real-time early warning system for sepsis based on a data integration system that can be implemented at the bedside of the intensive care unit (ICU), named SEPRES.

**Methods:** Data is collected from bedside devices through the integration hub and uploaded to the integration system through the local area network. The data integration system was designed to integrate vital signs data, laboratory data, ventilator data, demographic data, pharmacy data, nursing data, etc. from multiple medical devices and systems. It integrates, standardizes, and stores information, making the real-time inference of the early warning module possible. The built-in sepsis early warning module can detect the onset of sepsis within 5 hours preceding at most.

**Results:** Our data integration system has already been deployed in Ruijin Hospital, confirming the effectiveness of our system.

**Conclusions:** We highlight that SEPRES has the potential to improve ICU management by helping medical practitioners identify at-sepsis-risk patients and prepare for timely diagnosis and intervention.

## 1. Introduction

Sepsis is a syndrome of physiologic, pathologic, and biochemical abnormalities induced by infection [1]. It is a global medical problem associated with unacceptably high mortality, long-term morbidity, and a major cost burden on healthcare resources [1,3]. Early detection and timely administration of appropriate antibiotics are probably the most important factors in improving the prognosis of septic patients [4]. However, non-specific symptoms in septic patients lead to delayed diagnosis and delayed intervention [5].

Machine learning, including regression models, survival models, decision trees, and neural networks, has become a promising tool for predicting sepsis based on electronic medical records, laboratory data, and biomedical signals [6-12]. In 2016, Singer et al proposed a new definition (Sepsis-3) of sepsis, which defined sepsis as life-threatening organ dysfunction caused by a dysregulated host response to infection [1]. According to this, many recent papers defined sepsis by Sequential Organ Failure Assessment (SOFA) and infection instead of SIRS [13-19].

Most studies on sepsis prediction used historical medical data, such as the Medical Information Mart for Intensive Care (MIMIC) [20]. However, the raw data needed for clinical model inference, such as bedside data, laboratory data, demographic data, and doctor’s orders, usually come from different devices. Moreover, the information cannot interact directly due to differences in the data transfer protocols between devices. Data displayed on discrete devices can divide medical practitioners’ attention and hinder further data utilization. Efforts have been made to integrate bedside medical devices. Smielewski et al developed ICM+ software that allowed easy configuration and real-time trending of complex parameters derived from multiple bedside monitoring devices [21]. Meyer et al implemented a system for the operating room that integrates data from surgical and anesthesia devices, information systems, and a location tracking system [22]. Goldstein et al developed a real-time, physiologic data acquisition system in the pediatric intensive care unit [23]. Gjermundrod et al implemented the Intensive Care Window which can retrieve and integrate data from different patient monitoring devices in ICU [24]. Sun et al proposed an integrated system INSMA, which supports multimodal data acquisition, parsing, real-time data analysis, and visualization in the ICU [25]. In [26-28], the authors combined information systems with sepsis prediction, with data autonomously obtained from the electronic health record (EHR), and demonstrated the effectiveness of machine learning algorithms in predicting sepsis clinically. However, these data integration systems or prediction systems integrate a more limited number of devices and data types to present the complete perspective of a doctor. We aim to integrate various devices and systems at the bedside in the ICU to obtain high-density raw data and achieve real-time predicting of sepsis.

In this study, we developed a data integration system that integrates IntelliVue Information Center, Ventilators, Philips ICCA system, Laboratory Information System (LIS), and Hospital Information System (HIS), and established a real-time early warning system for sepsis in the ICU, named SEpsis PREdiction System (SEPRES).

## 2. Method

SEPRES comprises a data integration system with a sepsis early warning module. The data integration system completes the collection, storage, processing, and display of medical data. The sepsis early warning module comprises a sepsis prediction model and an interpretative tool. The sepsis prediction model is an ensemble model of gradient boosting machine and multilayer perceptron that can output the risk value of sepsis onset within 5 hours preceding at most. These models were first trained on the open-source dataset Medical Information Mart for Intensive Care (MIMIC-III) and then transferred to the private target hospital (Ruijin Hospital) dataset. Finally, the models were ensembled into the sepsis prediction model. The interpretative tool supplies information about how the model works by attributing importance value to each input feature. More information about the sepsis early warning module can be found in [29]. The study protocol complies with the Declaration of Helsinki, as revised in 2013, and was approved by the Ruijin Hospital Ethics Committee (No. 2020 [140]).

### 2.1. System Framework

As shown in Fig. 1, the system comprises a physical server with the PostgreSQL database to store the sepsis warning data and the webserver to deploy the portal for user access. The Web release system of the sepsis early warning system applies Brower/Server (B/S) architecture. The whole architecture can be divided into the following parts.

**Fig. 1.**
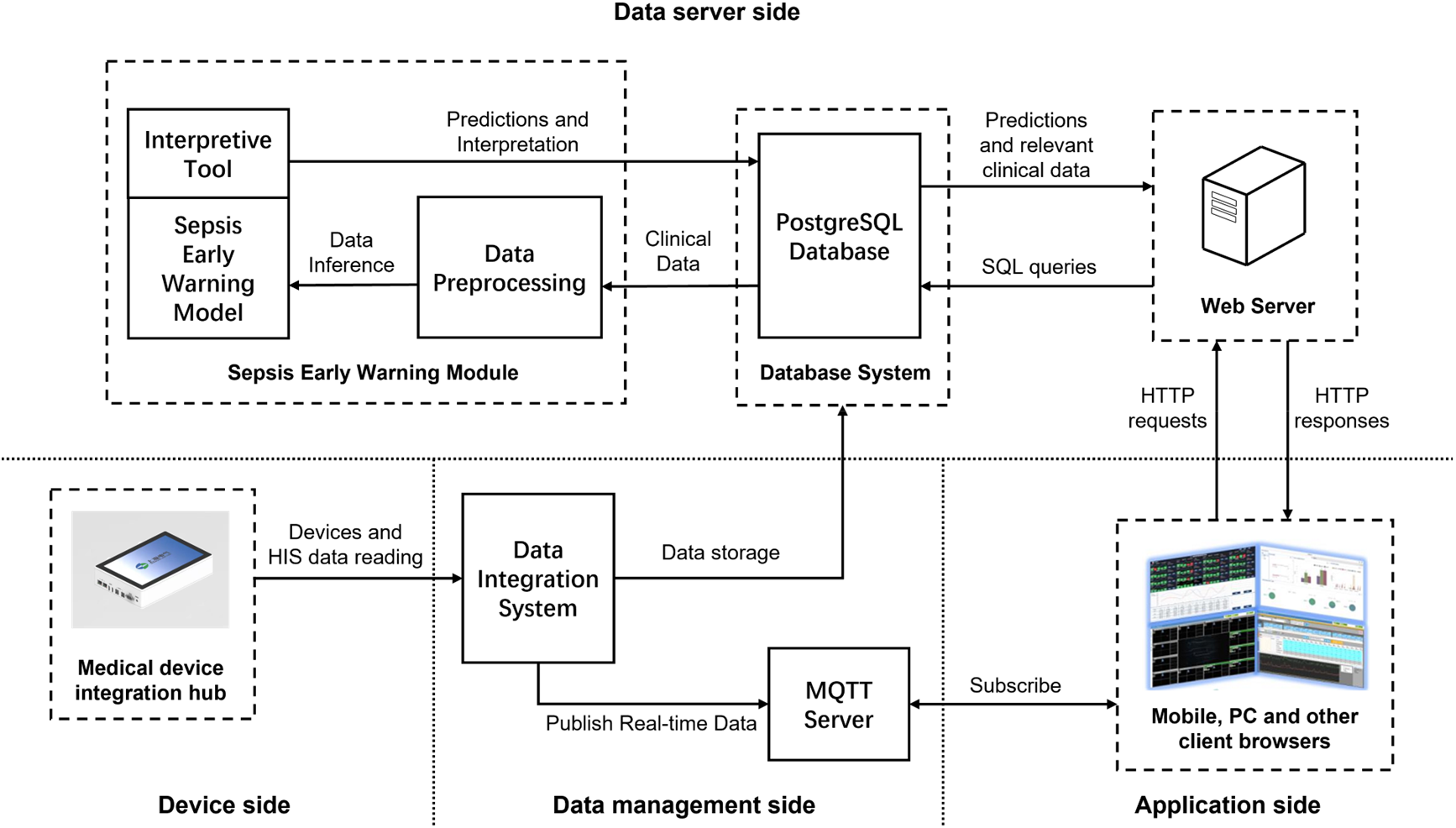
System deployment framework.

- Device side: The medical device integration hub transmits the device data to the data integration system through the local area network.
- Data management side: Heterogeneous data are integrated into the data integration system. The interface data, service data, and model predictions are stored and managed by the Structured Query Language (SQL) server, while the parts needed for the sepsis early warning module are sent to the PostgreSQL database. The Message Queuing Telemetry Transport (MQTT) server sends real-time data from the data integration system to the browser.
- Data server side: The web server uses the AJAX interface to respond to the browser’s request and calls the sepsis early warning module. Data fetching, data cleaning, feature extraction, standardization, and other preprocessing are implemented in turn. Model inference is then executed, and the predictions are stored in the PostgreSQL database. The data server side provides business support for the browser-side interface, including some related services (real-time calculation of the SOFA score, determination of suspected infection, data statistics, data charts, and historical data query).
- Application side: The user’s request is passed to the webserver in this layer, and the processing results are displayed in the system. The Java Script program is used for dynamic HTML page development, and the AJAX interface is used for data interaction with the webserver. Spring MVC is used to build full-featured MVC modules for web applications, combined with NODEJS to provide an elegant and highly maintainable method for creating templates. Users can use the system anytime and anywhere with a browser in various ways, such as on PCs and mobile terminals.

The data collected by SEPRES is shown in Table I. Considering data availability and importance, we extracted 63 variables for predicting sepsis in Ruijin Hospital, as shown in Appendix I.

**Table I.** The types of data collected from the devices and systems.

| Source device/system | Data type | Output medium | Format |
| --- | --- | --- | --- |
| IntelliVue Information Center | Vital signs data | Network | HL7 |
| PB 840 Ventilator | Ventilator data | RS-232 | According to the device output format |
| Maquet Servo-i Ventilator |  |  |  |
| Maquet Servo-s Ventilator |  |  |  |
| Philips ICCA | Pharmacy data, GCS, and urine output | Network | Web Services |
| Laboratory Information System | Laboratory data | Network | Web Services |
| Hospital Information System | Admission, discharge, and hospitalization data | Network | Web Services |

The data integration system queries the devices and systems at regular intervals or receives data sent by the system at regular intervals. The detailed modes and frequencies are shown in Table II.

**Table II.** The types of data collected from the devices and systems.

| Source device/system | Update time | Description |
| --- | --- | --- |
| IntelliVue Information Center | Per-minute | The IIC sends data every minute and the integration system receives |
| PB 840 Ventilator | Per-minute | The medical device integration hub queries data to the devices every minute and uploads it to the integration system |
| Maquet Servo-i Ventilator |  |  |
| Maquet Servo-s Ventilator |  |  |
| Philips ICCA | Recorded by nurses at regular intervals or when executing medical orders | The integration system regularly queries the ICCA database for recently updated nurse records and medical orders |
| Laboratory Information System | Recorded when reports are complete | The integration system regularly queries the LIS for completed inspection reports |
| Hospital Information System | Recorded when the patient is admitted to or leaves the ward | The integrated system regularly queries the HIS for the current status of patients in the ward |

### 2.2. Medical device integration hub

We developed a medical device integration hub that can acquire and transmit data from different brands of medical devices. The medical device integration hub consists of customized device connection lines, a hub, and an integrated data receiver. The identification module containing encoding is inserted into each medical device, enabling the hub to identify the type of online device and collect data automatically according to the communication protocol. The integrated data receiver receives and translates the raw data and uploads them to the integration server through the local area network. The medical device integration hub has the following functions:

- Device online services: detecting device connections and starting a data reading program corresponding to the device.
- Decoding: parsing raw data into structured data for further processing.
- Storage: storing parsed data into native memory.
- Remote Settings: supporting remote system setup and sending system status.
- Uploading: uploading the received data to the specified database.

Most medical devices communicate through the network or RS-232 port, and the device data are transmitted to the data integration system through the HL7 interface, Web Services, or other protocols.

## 3. Results

Our machine learning model obtained an area under the receiver operating characteristic curves (AUC) of 0.98 on MIMIC-III from 1-5h preceding, which outperformed most of the similar literature. As a real-world study of our system, we have deployed SEPRES in the ICU of Ruijin Hospital since February 2021, and collected the results of the system from February 2021 to June 2021, involving a total of 67 patients. No patients were excluded during this period. The AUCs were 0.94-0.94 and 0.86-0.9 respectively on the historical dataset of Ruijin Hospital and Ruijin real-world study from 1-5h preceding. The details can be found in [29]. We next focus on the implementation of the data integration system.

### 3.1. Model inference

We use Python to implement these models. We apply Python.Net package to realize the interaction between .NET Framework and Python, and SQLAlchemy package to realize the interaction between Python and the database system. When executing model inference, the following steps are performed sequentially:

1. Use SQL query statements to obtain the real-time features of patients, and pass them into Python through the interaction between Python and PostgreSQL.
2. Standardize the features by calling the scaler which is the standardizing function obtained in the training set.
3. Call the trained models to get the prediction results.
4. Call the interpretive tools to get the importance of the features based on the prediction results.
5. Transmit the results to the .NET Framework using Python.Net.
6. Output and store the prediction results and interpretations in a standard format.

### 3.2. System Deployment

Fig. 2 shows the medical device integration hub installed at Ruijin Hospital. The hub was placed at the bedside, receiving data from multiple devices via different interfaces shown at the bottom of the figure, storing the past 72 hours of data into native memory, and transmitting data with a time delay of under 10 s. The interfaces distributed on two sides of the hub include two universal network interfaces, four USB interfaces for mouse, keyboard, and U disk, two HDMI for extended display, one RS-232, and eight or sixteen USB and Ethernet multiplexing interfaces for medical devices. The hub can integrate data from the monitor, ventilator, infusion pump, and dialysis machine. These processed data were then transmitted to the data integration system.

**Fig. 2.**
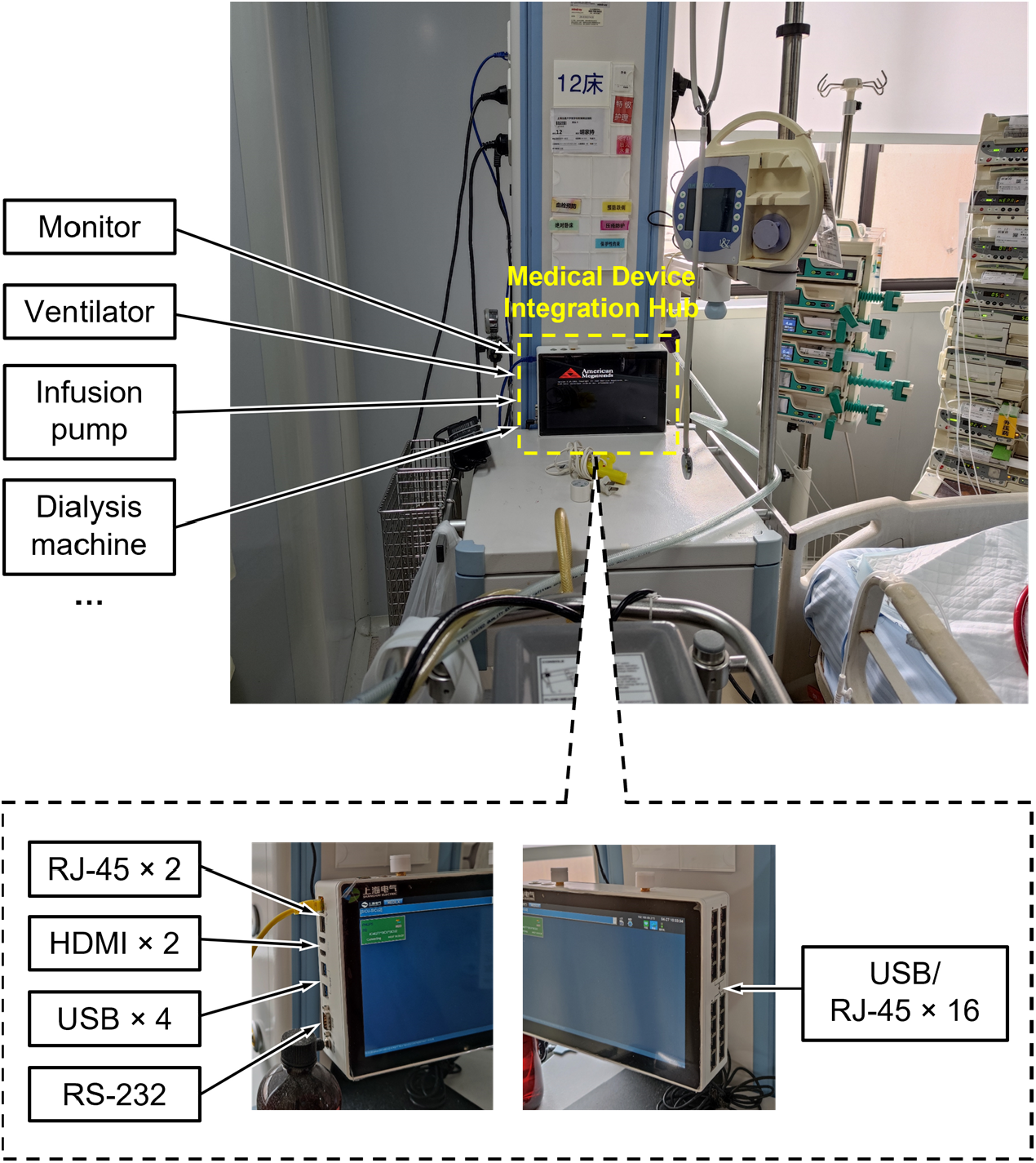
The medical device integration hub installed at Ruijin Hospital.

### 3.3. Patient statistics

We extracted the variable statistics on admission to the ICU collected by SEPRES for 67 patients, as shown in Table 3. The Missing column indicates the number of patients for which the variable was not recorded. The absence of records, especially laboratory variables, was attributed mainly to the short stay in the ICU for some patients, resulting in the lack of corresponding tests.

**Table III.** Variable statistics on admission.

| Variable | Number (%) | Missing | Variable | Number (%) | Missing |
| --- | --- | --- | --- | --- | --- |
| Gender-male | 43 (65.15) | 1 | Ventilation | 33 (50) | 1 |
|  | Mean (SD) | Missing |  | Mean (SD) | Missing |
| SOFA | 4.64 (3.37) | 0 | Platelets ($10^{12}/L$) | 180.02 (118.32) | 2 |
| Age (year) | 63.36 (17.13) | 1 | BUN (mg/dL) | 9.99 (8.38) | 2 |
| Weight (kg) | 63.54 (14.29) | 3 | Creatinine (mg/dL) | 1.7 (2.32) | 2 |
| Heart rate (bpm) | 88.83 (23.33) | 2 | Uric acid (mg/dL) | 4.87 (2.57) | 2 |
| Systolic blood pressure (mmHg) | 130.75 (27.63) | 2 | LDH (IU/L) | 505.29 (1849.89) | 2 |
| Diastolic blood pressure (mmHg) | 75.92 (18.54) | 2 | ALP (U/L) | 92.48 (59.21) | 2 |
| Mean arterial pressure (mmHg) | 93.18 (17.91) | 2 | AST (U/L) | 177.83 (851.23) | 2 |
| Respiratory rate (insp/min) | 19.74 (5.34) | 2 | Bilirubin (mg/dL) | 1.61 (1.65) | 2 |
| Temperature (Celsius) | 37.03 (0.79) | 2 | Bilirubin direct (mg/dL) | 10.98 (17.33) | 2 |
| SpO2 (%) | 98.44 (4.31) | 19 | Albumin (g/dL) | 3.03 (0.51) | 2 |
| pH | 7.4 (0.07) | 2 | Partial thromboplastin time (sec) | 32.89 (13.33) | 2 |
| PaO2 (mmHg) | 139.73 (65.91) | 2 | Prothrombin time (sec) | 14.72 (3.53) | 2 |
| SaO2 (%) | 99.04 (1.27) | 2 | INR | 1.26 (0.32) | 2 |
| AaDO2 (mmHg) | 129.86 (84.33) | 2 | Fibrinogen (mg/dL) | 339.38 (120.92) | 2 |
| PCO2 (mmHg) | 41.09 (14.31) | 2 | Lactate (mmol/L) | 2.25 (1.64) | 3 |
| Bicarbonate (mEq/L) | 23.94 (5.31) | 2 | Glucose (mg/dL) | 193.65 (87.6) | 2 |
| Base Excess | -0.13 (5.52) | 2 | Sodium (mEq/L) | 138.98 (6.7) | 2 |
| White blood cell count ($10^{12}/L$) | 10.32 (6) | 2 | Chloride (mEq/L) | 106.32 (6.35) | 2 |
| Neutrophils (%) | 85.85 (12.78) | 2 | Potassium (mEq/L) | 3.92 (0.63) | 2 |
| Monocytes (%) | 3.43 (2.68) | 2 | Phosphorus (mEq/L) | 3.37 (1.76) | 2 |
| Lymphocytes (%) | 8.31 (5.24) | 2 | Magnesium (mg/dL) | 2.05 (0.39) | 16 |
| Red blood cell count ($10^{12}/L$) | 5.1 (15.13) | 2 | Troponin I (ng/mL) | 0.25 (0.68) | 7 |
| Hemoglobin (g/dL) | 9.74 (2.55) | 3 | Creatine Kinase (IU/L) | 175.38 (254.81) | 1 |
| Hematocrit (%) | 30.97 (7.68) | 3 | Creatine Kinase MB (ng/mL) | 3.8 (6.12) | 1 |

### 3.4. System Operation

SEPRES provides predictions and explanations for every patient in the ICU every hour, including the risk of sepsis onset in the next 5 h, the influence of features on the predictions calculated by Shapley additive explanation (SHAP) [30], and SOFA predictions. SEPRES helps doctors focus on patients who are more likely to develop sepsis and observe changes in physiological data and conditions more conveniently.

The PC terminal of the user interface is deployed in the ICU of Ruijin Hospital. Fig. 3 shows an example of the page displaying all 12 patients in the ICU. Each patient takes a panel, and the title bar provides the patient’s ward number, patient identification number, name, and gender information. For privacy reasons, the image has been processed and the patient identifier has been removed. Recent trends in SOFA are shown in the upper left, and the maximum and minimum values of SOFA and sepsis-onset prediction in the last 24 hours are summarized in the upper right. The lower part of the panel shows the predicted sepsis-onset risk for the last 5 hours and the next 4 hours, with the trend fitted. High and low risks are shown by red and blue bars, respectively. By double-clicking on any panel, the influence of features calculated by SHAP for two models’ predictions are displayed, and the features with the highest absolute value of importance are displayed on the right side, as shown in Fig. 4. The title bar of this page provides information on the patient identification number, calculation time, and prediction time.

**Fig. 3.**
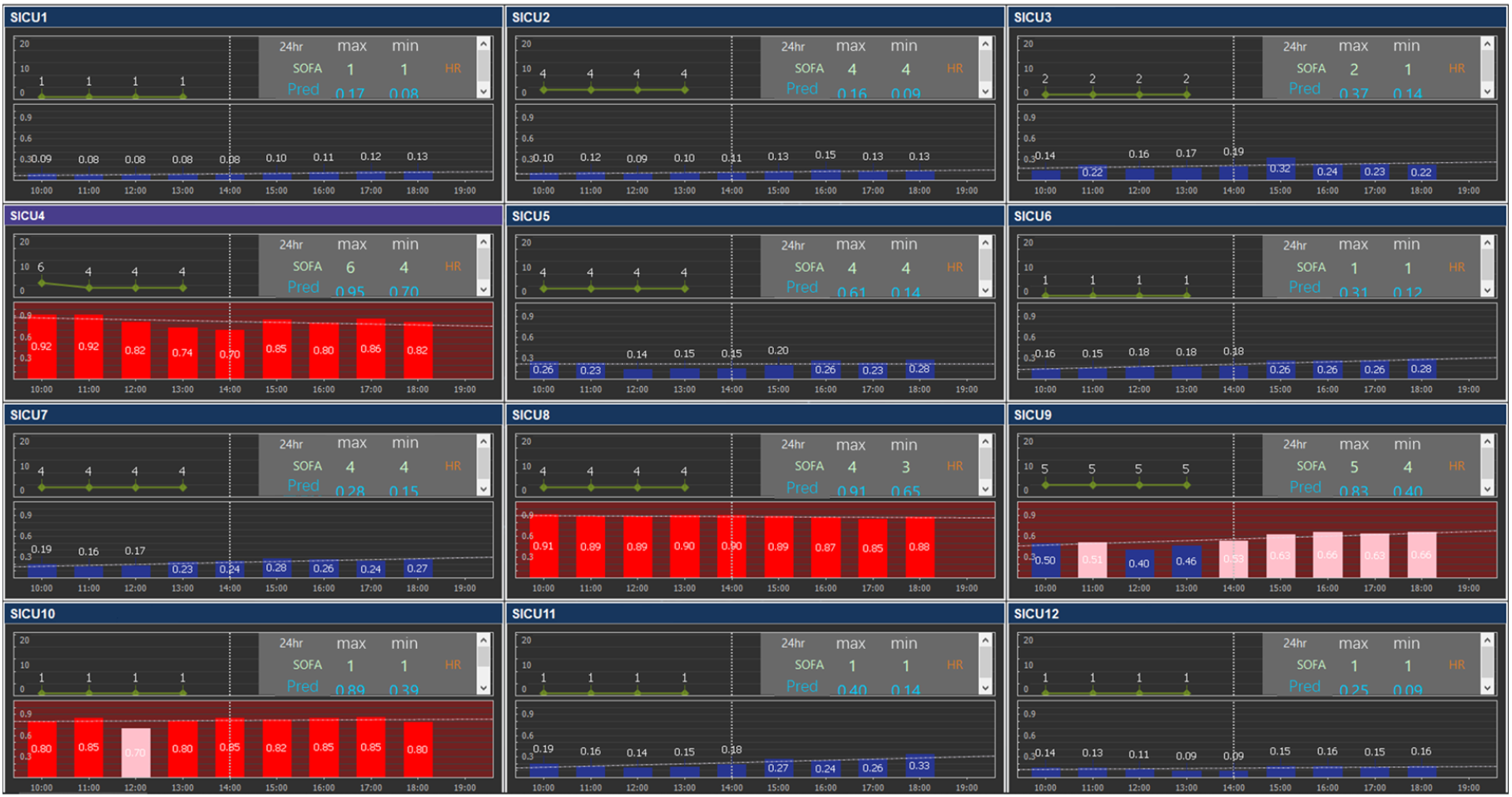
An example of the user interface. The original figure has been translated and the patient identifying information has been removed.

**Fig. 4.**
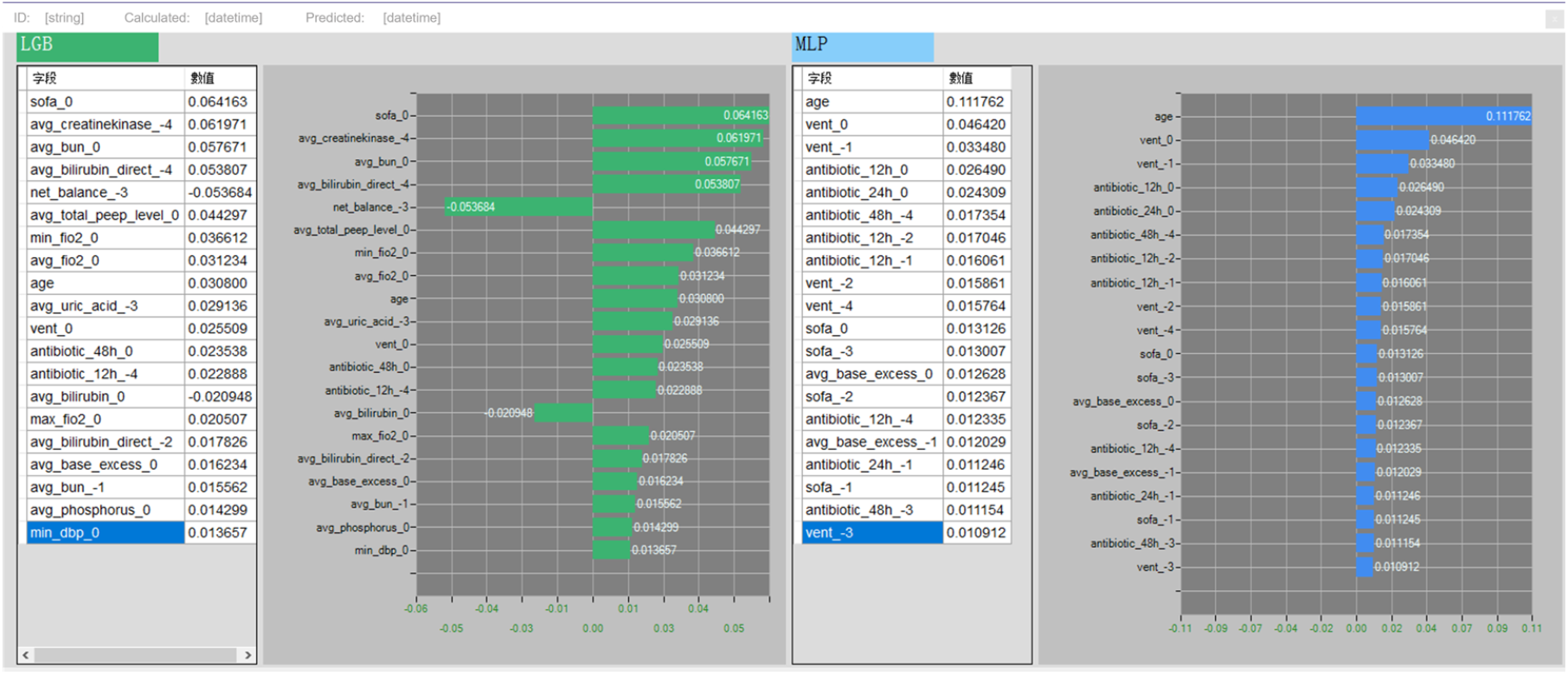
The example of SHAP values calculated for two models in a single prediction. The original figure has been translated and the patient identifying information has been removed.

Another major interface of the early warning system is shown in Fig. 5. In this interface, variable data for a specific patient during a certain historical period can be queried. On the top of the page is the filtering criteria, including ward number, patient identification number, query start time, and query end time. On the left side is the optional variable type. After the selection is completed, the results are displayed on the right side of the page, with numerical data and line graphs sorted by time on the top and bottom, respectively. Medical practitioners can filter variables of interest freely from any period in the past to track the patient’s condition promptly.

**Fig. 5.**
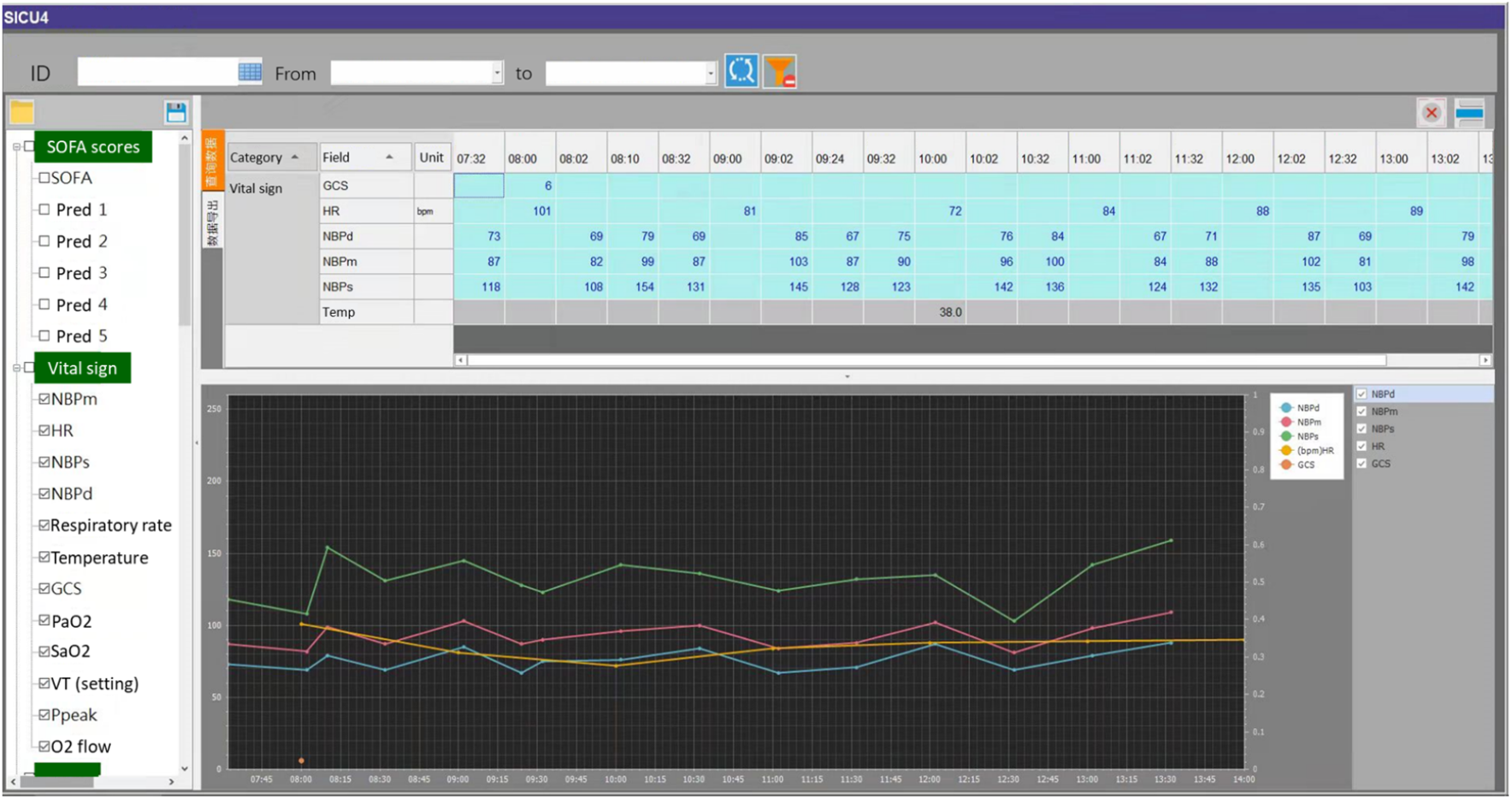
Historical data review for an individual patient. The original figure has been translated and the patient identifying information has been removed.

## 4. Discussion

Different types of monitoring equipment used in ICU often reflect various aspects of a patient’s status. These devices contain a large amount of information, however, due to the different data types and transmission protocols between different types and brands of devices, the data usually cannot interact with each other, creating a silo effect and making it difficult for further data utilization. Our system identified devices through customized modules, collected data according to the corresponding communication protocols, and then integrated them into one system, enabling high-density and real-time data recording. Vital signs and ventilation data are updated in SEPRES every minute and other data are updated at regular intervals, making real-time sepsis prediction possible. Although prediction was performed at an hourly frequency in our system, we highlight that a higher frequency of prediction is also feasible.

Data such as vital signs are generally of high resolution, which can lead to a lack of storage space. To solve this problem, we divided the data storage into three levels: at the device side, the data collected in real-time is stored in the integration hub, indexed by the device ID and time, stored in JSON format through NOSQL; at the central station side, the data from each integration hub is indexed by the device ID, patient ID and time, and stored in JSON format through NOSQL technology; at the server side, the data from the integration hubs are uploaded individually to the remote database host, and stored in different tables in the relational database according to different device types and data requirements. Each side can remove the obsolete data according to the time.

With access to rich data, we can leverage the data to perform higher-level tasks. In SEPRES, we used machine learning models to predict the onset of sepsis in real-time, giving risk predictions for each patient. Machine learning methods have been shown to be a promising approach for sepsis early warning [6-19,26-28]. Our results also confirm the feasibility of this approach. Furthermore, this workflow applies to alerts for other diseases. By utilizing the data integration system to collect the features and data required for model construction, we can conveniently construct different models for multiple tasks, such as disseminated intravascular coagulation and acute kidney injury.

In addition to performing basic ICU information system and real-time prediction functions, SEPRES can also provide a valuable source of data for future research works, including retraining of current models, other prediction tasks, data analysis, causal inference, etc.

## 5. Conclusion

In conclusion, we established an ICU bedside sepsis early warning system, SEPRES to achieve the real-time prediction of ICU patients through the data integration system. This system has been installed in Ruijin Hospital. Our real-world study confirms the feasibility of our system. The system can display the patient’s historical data in the user interface, to facilitate doctors to obtain the change of the patient’s condition intuitively. The risk of sepsis occurrence calculated by SEPRES allows medical practitioners to focus more on specific patients, enabling early diagnosis of sepsis and more effective management of ICU patients.

## Data Availability

All data produced in the present work are contained in the manuscript

## Acknowledgments

The authors thank the MIMIC team for their efforts in collecting and making the MIMIC data publicly available.

## Authors’ contributions

**QC:** Data Curation, Formal analysis, Software, Validation, Writing - Original Draft **RL:** Data Curation, Formal analysis, Validation, Writing - Original Draft **Lin C:** Conceptualization, Funding acquisition, Methodology, Supervision **Lai C:** Data Curation, Software, Validation **YH:** Resources **WL:** Funding acquisition, Project administration, Supervision, Writing - Review & Editing **LL:** Conceptualization, Formal analysis, Project administration, Resources, Supervision, Validation, Writing - Review & Editing

## Appendix I Complete list of variables used on Ruijin Hospital

We consulted the literature on predicting sepsis or SOFA scores and the variables mentioned in the literature and could be extracted from Ruijin Hospital dataset were selected, for a total of 63 variables [1-4].

Classified by the devices and systems, the 63 variables collected were:

**IntelliVue Information Center (Vital signs data):** MAP, heart rate, SBP, DBP, respiratory rate, temperature, PaO2, FiO2, SpO2.

**Ventilator data:** tidal volume, peak inspiratory pressure, total PEEP level, O2 flow rate, ventilation.

**ICCA (Pharmacy data, GCS, and urine output):** urine output in the past 24 hours, number of antibiotics in the past 12, 24, and 48 hours, rate of norepinephrine, epinephrine, dopamine, and dobutamine.

**Laboratory Information System (Laboratory data):** WBC, hemoglobin, hematocrit, creatinine, bilirubin, bilirubin direct, platelets, INR, PTT, AST, lactate, glucose, potassium, BUN, phosphorus, magnesium, chloride, troponin I, fibrinogen, PH, PCO2, bicarbonate, base excess, SaO2, albumin, PT, sodium, creatine kinase, creatine kinase-MB, LDH, ALP, uric acid, monocytes, lymphocytes, AaDO2, RBC, neutrophils.

**Hospital Information System:** weight, net balance, age.

**Calculated:** SOFA.

